# Fibrosis status, extra-hepatic multimorbidity and all-cause mortality in 53093 women and 74377 men with MASLD in UK Biobank

**DOI:** 10.1101/2025.03.09.25323329

**Authors:** Qi Feng, Chioma N Izzi-Engbeaya, Pinelopi Manousou, Mark Woodward

## Abstract

**Introduction:** People with MASLD had higher risk of extrahepatic multimorbidity, and fibrosis is the strongest prognostic factor for mortality in MASLD. This study aimed to investigate how fibrosis was associated with multimorbidity and their relationships with all-cause mortality.

**Methods:** We utilized data from the UK Biobank. MASLD was identified via a Fatty Liver Index ≥ 60 and the presence of cardiometabolic risk factors. The fibrosis-4 (FIB4) score was used to measure liver fibrosis. Multimorbidity was defined as having two or more long-term conditions (LTCs) from a prespecified list of 47 LTCs. Logistic regressions estimated the cross-sectional association between FIB4 scores and multimorbidity prevalence, while Cox models assessed the prospective association between FIB4 scores, multimorbidity and mortality.

**Results:** We included 127470 participants with MASLD (41.7% female, age 57.4 years, 21.3% with multimorbidity, 2.2% with high FIB4 scores). 14471 deaths were recorded during 13-year follow-up. Compared to low FIB4 scores, high FIB4 scores were associated with 41% higher prevalence of multimorbidity (OR 1.41 (95%CI 1.30-1.54)), and 94% higher all-cause mortality (HR 1.94 (95%CI 1.77-2.13)), with multimorbidity explaining 10% of this association, primarily driven by contributions from cardiovascular diseases, extrahepatic cancers, and chronic kidney disease.

**Conclusion:** FIB4 scores were positively associated with higher multimorbidity and mortality in MASLD patients. Multimorbidity explained 10% the relationship between fibrosis and mortality, with cardiovascular diseases, cancers and chronic kidney disease contributing notably to this mediation. These findings underscore the importance of managing both fibrosis and multimorbidity in MASLD patients.

## Introduction

Metabolic-dysfunction associated steatotic liver disease (MASLD), formerly known as non-alcoholic fatty liver disease (NAFLD), is the most common chronic liver disease, with an estimated global prevalence of 32.4% [1]. MASLD is linked to a range of hepatic and extra-hepatic diseases, particularly cardiovascular diseases (CVDs), and extrahepatic cancers [2,3], resulting in a higher risk of extrahepatic multimorbidity (defined as having two or more long-term conditions (LTCs)) [4].

Fibrosis status is the most important prognostic factor for severe liver diseases and mortality in people with MASLD [5]. A systematic review has shown that biopsy-confirmed advanced fibrosis stage increased mortality by 2.5 times, compared to early stage fibrosis [6]. Assessing liver fibrosis stages is therefore pivotal in MASLD management, with liver biopsy being the gold standard. However, liver biopsy is invasive, costly, requires significant clinical skills, carries extra risk of complications, and is maybe prone to sampling variability, thus making it impractical for large cohort studies. Consequently, various non-invasive assessments have been developed and adopted in international guidelines for MASLD management [7,8]. Among these, fibrosis-4 score (FIB4) has been validated and widely used, and linked to increased all-cause mortality [9,10]. Previous studied also reported FIB4 outperformed other non-invasive biomarker-based scores in predicting advanced fibrosis [11] and outperformed liver biopsy in prognosis prediction [12].

Currently, there has been a lack of evidence regarding the association between liver fibrosis and multimorbidity, and how their interplay is associated with all-cause mortality. Therefore, this study was aimed to investigate whether liver fibrosis status was associated with multimorbidity, and the potential role of multimorbidity in the association between fibrosis and all-cause mortality in people with MASLD.

## Methods

### Data and participants

UK Biobank is a prospective cohort of half million participants aged between 40 and 70 years old recruited across the UK between 2006 and 2010. During baseline assessment, data were collected on socioeconomic characteristics, lifestyle, health status, anthropometry and physical measures. The participants were followed up via linkage to national registries, and hospitalisation datasets. All participants provided informed consent.

We identified participants with MASLD using the definition proposed by Rinella M. et al. [13], considering liver steatosis, cardiometabolic risk factors and low alcohol consumption (< 20/30 g/day for female/male). We used a fatty liver index (FLI) ≥ 60 as a proxy for liver steatosis. FLI is a non-invasive biomarker for hepatic steatosis, calculated from body mass index (BMI), waist circumference, triglycerides, and gamma-glutamyl transferase (GGT) [14]. The cutoff value of 60 to define liver steatosis has been validated and used previously [15,16]. We considered the following five cardiometabolic risk factors: obesity (BMI ≥25 kg/m ^2^ AND/OR waist circumference > 94/80 cm (male/female)), diabetes (glycated haemoglobin (HbA1c)≥39 mmol/mol AND/OR diagnosis of type 2 diabetes AND/OR on treatment for type 2 diabetes), hypertension (systolic blood pressure (BP) ≥130 AND/OR diastolic BP ≥ 85 mmHg AND/OR on antihypertensive drug treatment or diagnosis of hypertension), high triglycerides (TG) (plasma TG ≥1.70 mmol/L AND/OR on lipid lowering treatment), and low high-density lipoprotein (HDL) cholesterol (HDL-cholesterol ≤1.0/1.3 mmol/L (male/female)) AND/OR on lipid lowering treatment).

We excluded women who were pregnant at baseline, and people who had missing data for calculating FLI or defining MASLD. We further removed those with alcohol related liver disease (ALD), metabolic dysfunction alcohol related liver disease (MetALD), or other chronic liver disease (including viral hepatitis, liver fibrosis, liver cirrhosis, hepatocellular carcinoma, hemochromatosis, Wilson’s disease, biliary cirrhosis, autoimmune hepatitis, primary sclerosing cholangitis, toxic liver disease and Budd-Chiari syndrome). For ALD and MetALD, we used the definitions proposed by Rinella et al. [13], while other chronic liver disease was ascertained via self-report and hospitalisation data; the code lists for these conditions are presented in supplementary methods.

We used the FIB4 score, a non-invasive surrogate marker for fibrosis status, which required age, serum aspartate aminotransferase and alanine aminotransferase levels, and platelet count [17]. We used the cutoff values 1.30 and 2.67 to categorise low, intermediate and high levels of FIB4 score for people < 65 years old, and 2.00 and 2.67 for people ≥ 65 years old [18]. Among people with MASLD, we further excluded those who had missing FIB4 data.

### Variable measurement

Multimorbidity is defined as having two or more LTCs from a prespecified list of 47 LTCs. We used a modified list of LTCs identified in a three-round Delphi study of 25 public participants and 150 healthcare professionals (including clinicians, researchers and policy makers) in 2021 [19]. The criteria for including these LTCs included their impacts on risk of death, quality of life, frailty, physical disability, mental health and treatment burden. Since MASLD is our index disease, we define multimorbidity excluding MASLD and its associated CMRFs. Among the conditions identified in this Delphi study, we further removed two liver conditions (hepatocellular carcinoma and chronic liver disease), as well as post-acute covid-19 and chronic Lyme disease (supplementary method). The conditions covered extrahepatic solid organ cancers, haematological cancers, cardiovascular, metabolic/endocrine, respiratory, digestive, renal, mental/behavioural and congenital conditions. We used the self-reported data and hospitalisation records on and before baseline, for phenotyping multimorbidity.

The outcome of interest was all-cause mortality, confirmed via national death registry records. Participants were censored at the date of death or the last date of follow-up (30 November 2022), whichever occurred first.

Ethnicity was classified into White, Asian, Black, and mixed/others. The Townsend Deprivation Index is a postcode-derived measure used to designate socioeconomic status. Lifestyle factors considered were smoking status (current, previous and never smoker), alcohol consumption, and physical activity. Alcohol consumption was assessed via weekly or monthly consumptions of red wine, white wine, champagne, beer, spirits, fortified wine and other alcoholic drinks; the consumption was converted to standard UK alcohol units and grams and summed up to derive the average daily alcohol consumption (g/day) [20]. Physical activity level was categorized into low, moderate and high levels, based on the frequency, duration and intensity of their physical activities. Systolic and diastolic blood pressure were measured by trained staff. Blood biochemistry markers included TG, HDL cholesterol, HbA1c and platelet. For all the categorical covariates, answers of “unknown”, “do not know”, “prefer not to say” were combined into one “unknown” category.

### Statistical analysis

The baseline characteristics of participants were summarised using mean with standard deviation (SD) or median with interquartile range (IQR), as appropriate, and frequency with percentage, stratified by FIB4 score levels.

We used bar charts to demonstrate the distribution of the number of LTCs stratified by FIB4 score levels. We calculated the prevalence (per 1000) for each LTC and ranked them in people with low, intermediate and high FIB4 scores, for males and females separately. We fitted logistic regression, adjusted for sex, age, education and Townsend Deprivation Index (in fifths), to estimate prevalence odds ratios (ORs) and their 95% confidence intervals (CIs) relating cross-sectionally FIB4 score levels and prevalence of multimorbidity (defined as having ≥ 2 LTCs) at baseline. Similar logistic regression models were also fitted to estimate association between FIB4 score levels and each LTC; p values were corrected for multiple testing using false discovery rate (FDR) method [21]. We estimated the association between FIB4 score levels and multimorbidity in subgroups stratified by age (< 65, ≥ 65), Townsend Deprivation Index (fifths), education, smoking, physical activity level, and BMI levels (< 25, 25-30, ≥ 30).

Cox proportional hazard regression models were used to assess the prospective association between FIB4 score levels and all-cause mortality, expressed as hazard ratio (HR) and 95% CI, using a low FIB4 score level as reference. Models were stratified by region and age group, and adjusted for sex, ethnicity, education, Townsend Deprivation Index (in fifths), physical activity level, smoking status, and alcohol consumption. The proportional hazard assumption was examined by scaled Schoenfeld residuals, and no evidence was observed for its violation. To test whether the association of FIB4 score levels was independent of multimorbidity, we additionally adjusted for multimorbidity in the model to see whether it would change the coefficient estimates, and if yes, by how much; we calculated the reduction in coefficients before and after additional adjustment of multimorbidity, to quantify the percentage of the associations was accounted for by multimorbidity. Similarly, we also adjusted for each LTC one by one in the Cox model, and to see how much of the association was accounted for by each LTC.

We also calculated the multimorbidity-adjusted associations between FIB4 score levels and all-cause mortality in subgroups stratified by age, Townsend Deprivation Index, education, smoking status, physical activity level, BMI levels and multimorbidity status. For sensitivity analysis, we excluded the first two years of follow-up.

All analyses were conducted in R.

## Results

### Baseline characteristics

We included 127470 participants (41.7% females, mean age 57.4 years) (figure 1). Most of the participants had low FIB4 scores (70.6%), and 2.2% had high FIB4 scores. Compared to people with high FIB4 scores, those with lower FIB4 scores were more likely to be younger, female, better educated, physically inactive, and less likely to smoke or consume alcohol. They also had lower waist circumference, lower systolic blood pressure but higher diastolic blood pressure and platelet count. (table 1, supplementary table 1)

**Table 1:** Baseline characteristics of participants with baseline MASLD by FIB-4 score levels.

|  | FIB4 low<br>n = 90035 (70.6%) | FIB4 intermediate<br>n = 34668 (27.2%) | FIB4 high<br>n = 2766 (2.2%) | Overall<br>n = 127470 (100.0%) |
| --- | --- | --- | --- | --- |
| Sex, female | 41343 (45.9%) | 11001 (31.7%) | 748 (27.0%) | 53093 (41.7%) |
| Age, years | 56.6 (8.4) | 59.3 (5.6) | 62.9 (5.7) | 57.4 (7.9) |
| Townsend deprivation index,<br>1st fifth (least deprived) | 17698 (19.7%) | 7323 (21.1%) | 541 (19.6%) | 25562 (20.1%) |
| 5th fifth (most deprived) | 18328 (20.4%) | 6511 (18.8%) | 555 (20.1%) | 25394 (19.9%) |
| Education, higher education | 23365 (26.0%) | 9231 (26.6%) | 640 (23.1%) | 33236 (26.1%) |
| Ethnicity, white | 83828 (93.1%) | 32297 (93.2%) | 2579 (93.2%) | 118704 (93.1%) |
| Smoking, never | 48103 (53.4%) | 17831 (51.4%) | 1327 (48.0%) | 67261 (52.8%) |
| Alcohol drinking, g/day | 7.9 (8.8) | 9.3 (9.2) | 9.2 (9.2) | 8.3 (8.9) |
| Physical activity, high | 22043 (24.5%) | 9696 (28.0%) | 797 (28.8%) | 32536 (25.5%) |
| FIB4 score | 1.1 (0.3) | 1.7 (0.3) | 4.7 (20.3) | 1.3 (3.1) |
| ALT, log<sub>10</sub> U/L | 1.4 (0.2) | 1.4 (0.2) | 1.5 (0.3) | 1.4 (0.2) |
| AST, log<sub>10</sub> U/L | 1.4 (0.1) | 1.5 (0.1) | 1.6 (0.2) | 1.4 (0.1) |
| GGT, log<sub>10</sub> U/L | 1.6 (0.3) | 1.6 (0.3) | 1.7 (0.4) | 1.6 (0.3) |
| BMI, kg/m<sup>2</sup> | 31.9 (4.7) | 31.4 (4.4) | 31.5 (4.6) | 31.8 (4.6) |
| Waist circumference, cm | 102.4 (10.2) | 103.0 (10.1) | 104.2 (10.6) | 102.6 (10.2) |
| Waist-to-height ratio, % | 60.7 (6.2) | 60.2 (6.0) | 60.9 (6.4) | 60.6 (6.2) |
| Systolic blood pressure,<br>mmHg | 141.2 (17.4) | 142.5 (17.6) | 143.8 (18.6) | 141.6 (17.5) |
| Diastolic blood pressure,<br>mmHg | 85.2 (9.7) | 85.0 (9.8) | 83.8 (10.3) | 85.1 (9.7) |
| Triglycerides, mmol/L | 2.2 [1.3] | 2.1 [1.3] | 2.0 [1.3] | 2.1 [1.3] |
| HDL cholesterol, mmol/L | 1.1 (0.4) | 1.1 (0.4) | 1.1 (0.4) | 1.1 (0.4) |
| HbA1c, mmol/mol | 37.3 (10.1) | 36.8 (9.8) | 38.2 (11.9) | 37.2 (10.1) |
| Platelet, 10<sup>9</sup> cells/L | 274.7 (56.8) | 209.8 (38.7) | 149.6 (51.6) | 254.4 (61.8) |
| Hypertension | 72094 (80.1%) | 28023 (80.8%) | 2235 (80.8%) | 102352 (80.3%) |
| Obesity | 89089 (98.9%) | 34234 (98.7%) | 2706 (97.8%) | 126029 (98.9%) |
| Diabetes | 28339 (31.5%) | 10350 (29.9%) | 1047 (37.9%) | 39736 (31.2%) |
| High TG | 66192 (73.5%) | 24639 (71.1%) | 1788 (64.6%) | 92619 (72.7%) |
| Low HDL | 43112 (47.9%) | 14955 (43.1%) | 1323 (47.8%) | 59390 (46.6%) |

HbA1c: glycated hemoglobin. HDL: high density lipoprotein. Number(number): mean(SD). Number(percent%): frequency(percent). Number[number]: median[interquartile range]. FIB-4 score levels were defined using the cutoff values 1.30 and 2.67 to categorise low, intermediate and high levels for people < 65 years old, and 2.00 and 2.67 for people ≥ 65 years old.

**Figure 1:**
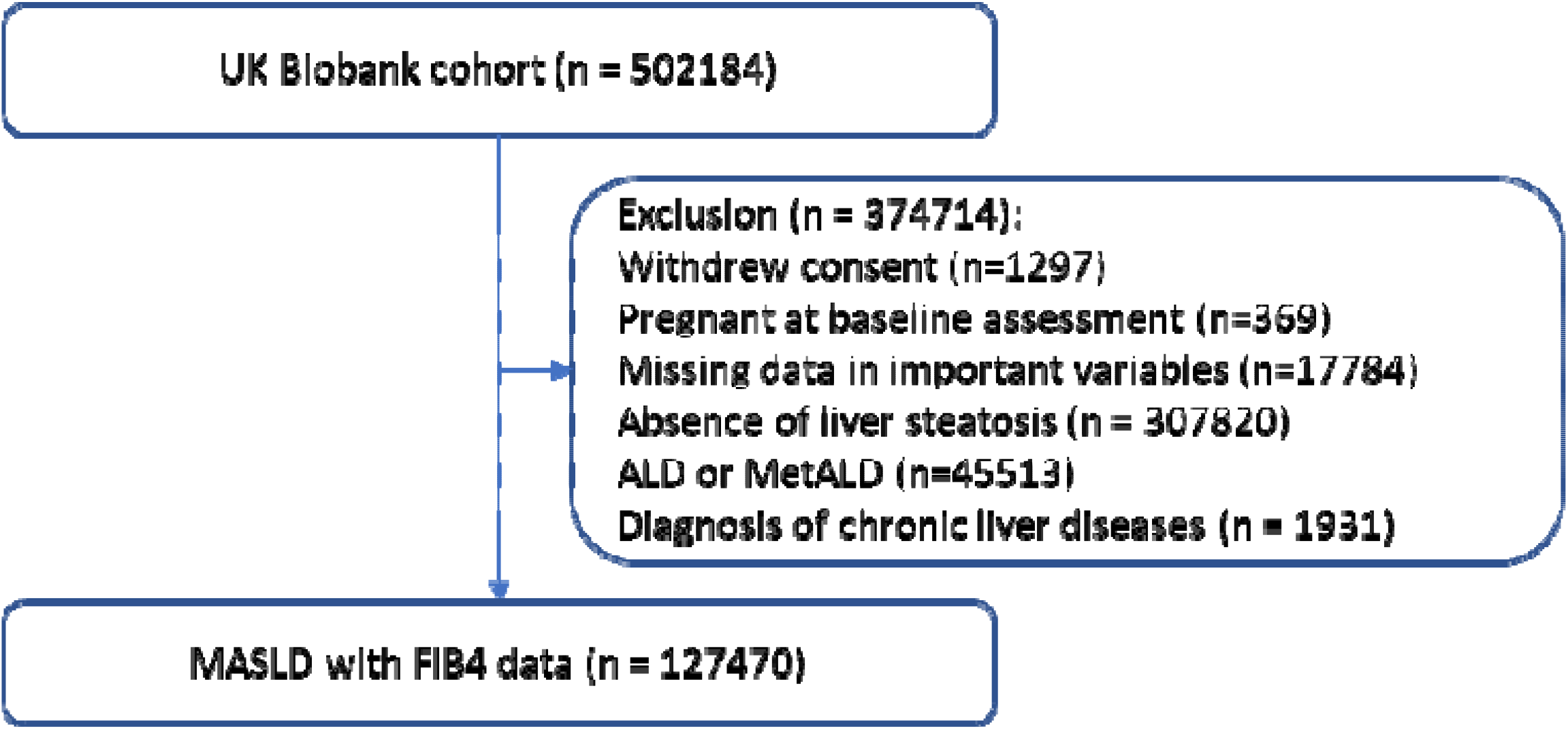
The flowchart of participant selection. MASLD: metabolic dysfunction-associated steatotic liver disease. ALD: alcohol related liver disease. MetALD: metabolic dysfunction and alcohol related liver disease. All data as recorded at baseline.

### FIB4 scores and multimorbidity

At baseline, 49.5% participants had no prevalent LTC, while 21.3% had prevalent multimorbidity (≥2 LTCs). The prevalence of multimorbidity was notably higher among people with high FIB4 scores (29.9%) than those with low (20.8%) or intermediate (21.8%) scores (figure 2). Multimorbidity was also more common in females than in males (25.0% vs. 18.6%), and in people ≥ 65 years than in people < 65 years (28.3% vs. 19.3%).

**Figure 2:**
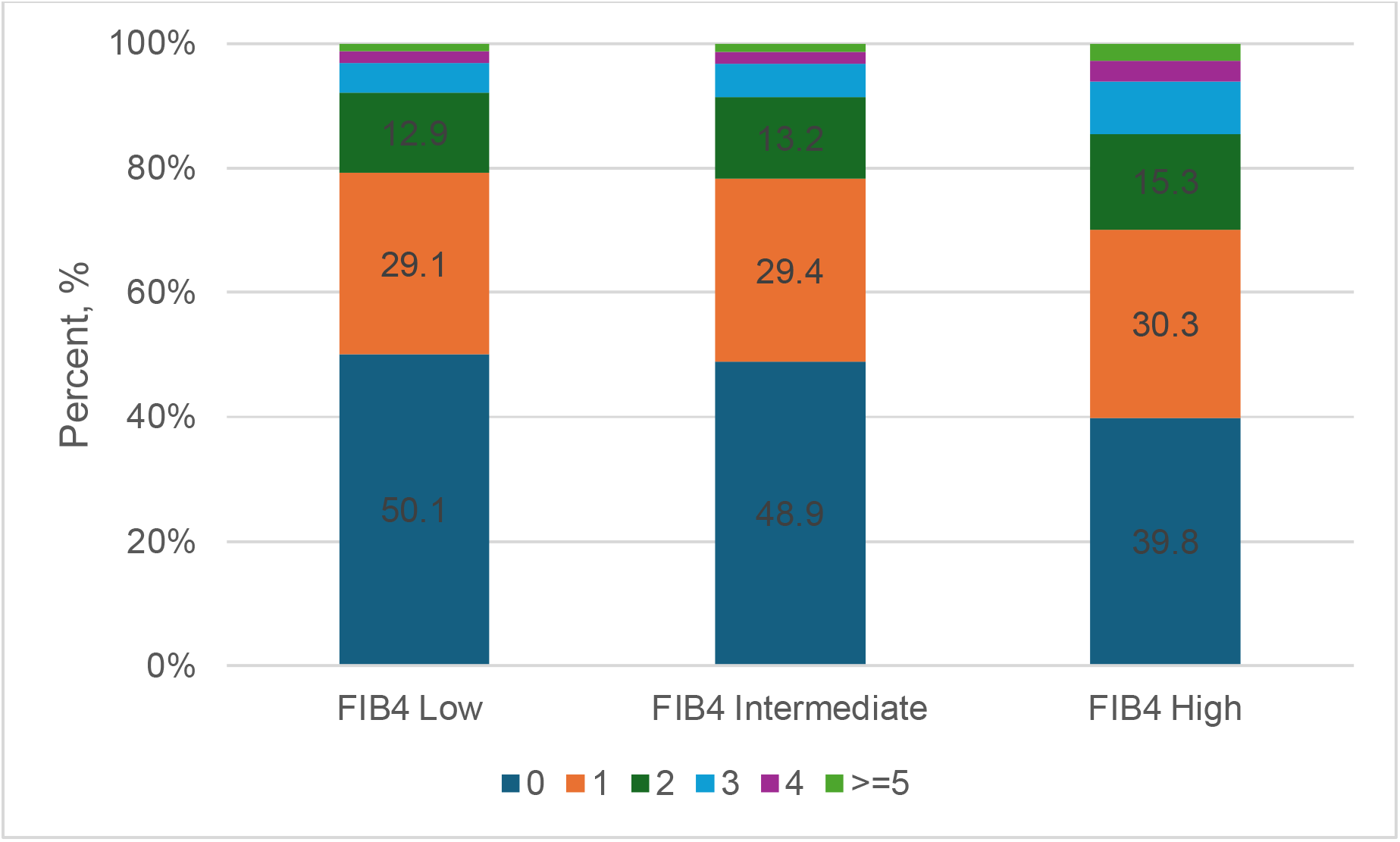
Bar chart of number of long-term conditions in people with MASLD by FIB4 score levels. FIB-4 score levels were defined using the cutoff values 1.30 and 2.67 to categorise low, intermediate and high levels for people < 65 years old, and 2.00 and 2.67 for people >= 65 years old.

After adjusting for sex, education and deprivation, compared to low FIB4 scores, high FIB4 scores were cross-sectionally associated with higher prevalence of multimorbidity, with ORs of 1.13 (1.09, 1.16) for intermediate and 1.69 (1.55, 1.84) for high FIB4 score levels. Further adjustment for age significantly reduced the associations, with ORs decreased to1.04 (1.01, 1.08)) and 1.41 (1.30, 1.54), respectively. These associations were stronger in males than in females (OR for high vs. low FIB4 scores: 1.42 (1.29, 1.58) and 1.26 (1.08, 1.47) for males and females, relative OR (male to female) 1.32 (1.09, 1.59)). Subgroup analyses stratified by age, Townsend Deprivation Index, education, smoking, physical activity and BMI, showed generally similar results (Supplementary table 2).

Supplementary figure 1 illustrates the relative ranks of LTCs by prevalence across low, intermediate and high FIB4 score levels. Supplementary table 3 provides the crude prevalences for these LTCs. Figure 3 and supplementary table 4 show the associations between FIB4 score levels and LTC prevalences, overall and by sex. Overall, higher FIB4 scores were associated with lower prevalences of asthma and chronic obstructive pulmonary disease (COPD), but higher prevalences of CVDs (e.g., ischemic heart disease (IHD), heart valve disorder, heart failure, arrythmia and aneurysm), solid organ cancers, haematological cancers, chronic kidney disease (CKD), mental/behavioural conditions (e.g., substance use disorder, schizophrenia and bipolar disorders), HIV/AIDS, thyroid disorders, gout, congenital diseases and anaemia. High FIB4 scores were additionally associated with venous thromboembolism and multiple sclerosis in males.

**Figure 3:**
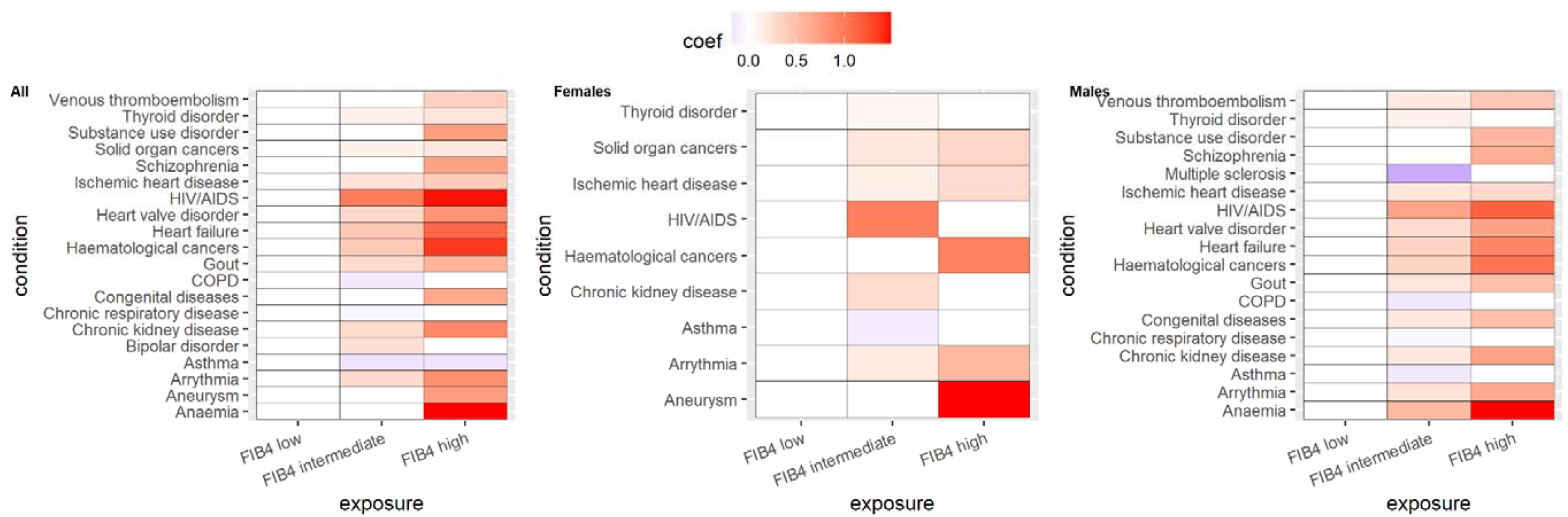
Heatmap showing the associations between baseline FIB-4 score levels and prevalent long-term conditions in people with MASLD. Coefficients were estimated using logistic regression, with low FIB-4 score level as reference. Red colour indicates higher prevalence, and blue lower prevalence. White colour indicates reference group (low FIB-4 scores) or insignificant differences. The figure shows only the long-term conditions that showed significant differences for intermediate or high FIB4 score levels. FIB-4 score levels were defined using the cutoff values 1.30 and 2.67 to categorise low, intermediate and high levels for people < 65 years old, and 2.00 and 2.67 for people >= 65 years old.

### FIB4 scores, multimorbidity and all-cause mortality

During a median follow-up of 13.0 years, 14471 deaths (9379 males) were recorded. Compared to people with low FIB4 scores, those with intermediate and higher scores had a 45% (HR (95%CI) 1.45 (1.38, 1.51)) and 94% (1.94 (1.77, 2.13)) higher all-cause mortality, respectively. Sensitivity analysis by excluding the first two years of follow-up showed similar results. When additionally adjusted for multimorbidity, the HR estimates reduced to 1.40 (1.34, 1.46) for intermediate and 1.82 (1.66, 2.00) for high FIB4 scores, respectively. This suggested that multimorbidity partly accounted for the association between FIB4 scores and mortality, by 9.0% and 10.0% for intermediate and high FIB4 levels, respectively.

The association was stronger in females than in males. For intermediate FIB4 scores, the HRs were 1.50 (1.37, 1.63) in females and 1.36 (1.29, 1.44) in males, with a relative HR (RHR, male to female) of 0.89 (0.82, 0.96). For high FIB4 scores, the HRs were 2.21 (1.81, 2.70) in females and 1.71 (1.54, 1.90) in males, with an RHR of 0.76 (0.64, 0.91). (table 3)

**Table 3:** Associations between baseline FIB4 score levels and all-cause mortality, with and without adjustment for multimorbidity.

|  |  | Events/total | HR (95%CI) * | HR (95%CI) with additional adjustment of multimorbidity | % reduction in coefficient estimates after adjustment** |
| --- | --- | --- | --- | --- | --- |
| Overall | FIB4 low | 9517 / 90035 | Reference | Reference |  |
|  | FIB4 intermediate | 4247 / 34669 | 1.45 (1.38, 1.51) | 1.40 (1.34, 1.46) | 9.0 |
|  | FIB4 high | 707 / 2766 | 1.94 (1.77, 2.13) | 1.82 (1.66, 2.00) | 10.0 |
| Females | FIB4 low | 3739 / 41343 | Reference | Reference |  |
|  | FIB4 intermediate | 1180 / 11002 | 1.53 (1.40, 1.66) | 1.50 (1.37, 1.63) | 4.5 |
|  | FIB4 high | 173 / 748 | 2.33 (1.91, 2.84) | 2.21 (1.81, 2.70) | 6.2 |
| Males | FIB4 low | 5778 / 48692 | Reference | Reference |  |
|  | FIB4 intermediate | 3067 / 23667 | 1.42 (1.35, 1.50) | 1.36 (1.29, 1.44) | 11.1 |
|  | FIB4 high | 534 / 2018 | 1.83 (1.65, 2.04) | 1.71 (1.54, 1.90) | 11.6 |

The Cox model was stratified by region and age group, adjusted for ethnicity, education, Townsend Deprivation Index (in fifths), physical activity, smoking status, and alcohol consumption. For the sex-mixed model, the model was additionally adjusted for sex. \*\*: calculated as the (unadjusted coefficient – adjusted coefficient)\*100/unadjusted coefficient. FIB-4 score levels were defined using the cutoff values 1.30 and 2.67 to categorise low, intermediate and high levels for people < 65 years old, and 2.00 and 2.67 for people ≥ 65 years old.

We assessed the linear association between FIB4 score and all-cause mortality in a subset of participants after removing those with potential outlier FIB4 scores (> 99.5% quantile, n = 641), and found that each 1 unit increase in FIB4 score was associated with 44% higher mortality (1.44 (1.39, 1.48)), more pronounced in females (1.52 (1.44, 1.61)) than in males (1.39 (1.34, 1.45)).

This multimorbidity-adjusted association between FIB4 scores and all-cause mortality remained consistent in subgroups stratified by age, Townsend Deprivation Index, education, smoking, physical activity and BMI levels, and multimorbidity status. (supplementary table 5)

Table 4 shows the results of how additional adjustment of each individual LTCs changed the association between FIB4 scores and all-cause mortality, for selected LTCs, while supplementary table 6 shows detailed results for all LTCs. The adjustment of CVDs (IHD, heart failure, stroke, heart valve disorder and arrythmia), solid organ cancers, haematological cancers, and CKD resulted in >1% reduction in coefficient estimates. IHD, heart failure and arrythmia demonstrated the largest mediation.

**Table 4:** associations between baseline FIB4 score level and all-cause mortality, with and without additional adjustment for long-term conditions (showing long term conditions with reduction in coefficient estimates > 1%).

| Long-term condition<br>additionally adjusted for | FIB4 Intermediate vs. low |  | FIB4 High vs. low |  |
| --- | --- | --- | --- | --- |
|  | HR (95%CI) | % reduction in<br>coefficient<br>estimates | HR (95%CI) | % reduction<br>in coefficient<br>estimates |
| Without adjustment for<br>long-term condition | 1.45 (1.38, 1.51) |  | 1.94 (1.77, 2.13) |  |
| **Additionally adjusted for**<br>**the condition:** |  |  |  |  |
| Ischemic heart disease | 1.40 (1.34, 1.46) | 9.5 | 1.85 (1.69, 2.03) | 7.1 |
| Heart failure | 1.42 (1.36, 1.49) | 4.2 | 1.87 (1.70, 2.05) | 5.8 |
| Stroke | 1.44 (1.38, 1.51) | 1.0 | 1.93 (1.76, 2.12) | 1.1 |
| Solid organ cancers | 1.43 (1.36, 1.49) | 4.0 | 1.92 (1.75, 2.11) | 1.5 |
| Haematological cancers | 1.44 (1.38, 1.51) | 0.9 | 1.91 (1.74, 2.10) | 2.5 |
| Chronic kidney disease | 1.44 (1.37, 1.50) | 1.7 | 1.92 (1.75, 2.10) | 2.0 |
| Heart valve disorder | 1.44 (1.38, 1.50) | 1.7 | 1.91 (1.74, 2.09) | 2.7 |
| Arrhythmia | 1.43 (1.36, 1.49) | 4.0 | 1.87 (1.70, 2.05) | 5.9 |

FIB-4 score levels were defined using the cutoff values 1.30 and 2.67 to categorise low, intermediate and high levels for people < 65 years old, and 2.00 and 2.67 for people ≥ 65 years old.

## Discussion

In this cohort of 127470 individuals with high risk of MASLD identified in UK Biobank, FIB4 scores were positively associated with the prevalence of multimorbidity, particularly with CVDs, extrahepatic cancers, CKD and mental/behavioural conditions. We observed a significant positive association between FIB4 scores and all-cause mortality, with multimorbidity accounting for 10% of such association, mainly driven by CVDs, cancers and CKD.

We found a positive link between FIB4 scores and prevalence of multimorbidity. A previous study reported that diabetic MASLD patients with advanced fibrosis stage (measured with transient elastography) had on average one more comorbidities (3.4 vs. 2.5), compared to those with lower fibrosis stages; however, this study had a small sample size of 82 individuals, considered only 15 LTCs to define multimorbidity, and only descriptively examined this association without adjusting for any confounders [22]. Our analysis highlighted the role of age in this association. Prior to adjusting for age, intermediate and high levels of FIB4 were associated with 13% and 69% higher risks of multimorbidity, respectively. However, adjusting for age reduced the associations: for intermediate FIB4 scores, the association became minimal, suggesting the association was mainly mediated by age; for high FIB4 scores, the OR reduced from 1.69 to 1.41, demonstrating age as a mediator. Among people ≥ 65 years old, both intermediate and high levels of FIB4 scores were associated with increased multimorbidity.

In our investigation on FIB4 scores and prevalence of specific LTCs, we found that higher FIB4 scores were positively associated with several conditions, including CVDs, solid organ cancers, haematological cancers, CKD, mental/behavioral diseases and thyroid disorder. The associations were observed in both males and females. While previous studies have reported associations between MASLD and these conditions [23–28], the relationships between fibrosis status and these conditions have been less clear. Prior research has indicated associations of liver fibrosis with CVDs [29] and CKD [30], but evidence regarding associations with extrahepatic cancers and mental/behavioral conditions remains inconclusive in people with MASLD [31].

The positive association between FIB4 scores and all-cause mortality in MASLD people was consistent with previous systematic reviews [6,32]. In contrast, some previous UK Biobank investigations reported null evidence for such associations. For instance, Roca-Fernandez A. et al. found no significant association between FIB4 scores and all-cause mortality (HR 0.96 (0.80, 1.15)) in 33616 UK Biobank participants with liver imaging data (27% with liver steatosis) with 2.5 year of follow-up [33]. Hydes T. et al. found that the association between FIB4 scores and mortality became null (HR 1.17 (0.94, 1.45)) after adjusting for smoking, diabetes, kidney function measures in 10512 people with both MASLD and CKD [34]. These discrepancies may be attributed to their small sample size, short follow-up periods and the inclusion of participants with specific conditions in these studies.

This association between FIB4 scores and all-cause mortality was more pronounced in females than in males, which was consistent with a previous study in the US [9]. A systematic review and meta-analysis also identified sex as a significant contributor in the meta-regression, suggesting different underlying associations between males and females [6]. This suggested potential sex-difference in MASLD prognosis, as suggested by previous research [35,36], and the observed sex dimorphism may possibly be attributable to sex hormone and lifestyle differences.

We also found that adjustment of multimorbidity reduced the associations between FIB4 and mortality, and multimorbidity was estimated to accounted for approximately 10% of the observed associations, higher in males than in females (11.1 % vs. 4.5% for intermediate FIB4 scores, 11.6% vs. 6.2% for high scores). Notably, this reduction was mainly driven by extrahepatic cancers, CKD and CVDs. Among these conditions, IHD, heart failure, and arrythmia contributed the most to the mediation. These conditions have been previously linked to MASLD [23,24,26].

Although monitoring fibrosis progression has been one of the key focus in MASLD management [37], our findings suggest that multimorbidity also plays a significant role and should be emphasized in the management of MASLD. The previously observed association between fibrosis and mortality has been found to be partly accounted for by multimorbidity, particularly by CVDs, extrahepatic cancers and CKD. This underscores the importance of a comprehensive approach that address both fibrosis and these comorbidities in disease management.

This study is the first large population-based cohort study to examine the relationship between liver fibrosis and multimorbidity, and it also provided evidence that multimorbidity partly accounted for the association between fibrosis and mortality. This study’s strengths include a large sample size, long follow-up time, detailed phenotyping of multimorbidity, and robust sensitivity and subgroup analyses. However, several limitations should be acknowledged. First, the study population was drawn from UK Biobank, which is predominantly white, less deprived, and healthier than the general UK population [38], therefore it may limit the generalisability of our results to other populations. Second, we used FLI to assess steatosis, instead of biopsy histological data or accurate imaging techniques. Nevertheless, FLI has been proven to be a valid surrogate for liver steatosis [16]. Third, we used FIB4 score as a non-invasive surrogate measurement for liver fibrosis stage, instead of the conventional gold standard of liver biopsy, which was also not available. FIB4 has been demonstrated to be an accurate surrogate for fibrosis staging [11]. Fourth, our analysis only considered multimorbidity measured at baseline, without capturing information on how the multimorbidity would change over time. Finally, we did not account for the temporal sequence of LTCs, and the relatively small number of prevalent cases for certain conditions may also limited our statistical power.

Our study highlighted that higher FIB4 scores, reflecting more advanced liver fibrosis in people with MASLD, were associated with higher prevalence of multimorbidity and higher all-cause mortality. Multimorbidity accounted for approximately 10% of the association between fibrosis and all-cause mortality, mainly driven by CVDs, extrahepatic cancer and CKD. We also found that the association between FIB4 scores and all-cause mortality was stronger in females compared to males, highlighting potential sex differences in MASLD prognosis.

## Data Availability

All data produced are available online at UK Biobank

## Statements

## Acknowledgement

We sincerely thank the UK Biobank participants and staff for their contribution to this valuable data resource. This study was conducted under application number 74018.

## Data sharing statement

UK Biobank data are available to registered researchers at https://www.ukbiobank.ac.uk/.

## Author contribution

QF conceived the research idea. QF conducted data analysis. All authors interpreted results. QF drafted the manuscript. All authors critically reviewed and revised the manuscript.

## Conflict of interests

None.

## Funding

This research/study/project was funded/supported by the NIHR Imperial Biomedical Research Centre (BRC) [NIHR203323]. The views expressed are those of the author(s) and not necessarily those of the NIHR or the Department of Health and Social Care.

The Section of Investigative Medicine and Endocrinology at Imperial College London is funded by grants from the MRC, NIHR and is supported by the NIHR Biomedical Research Centre Funding Scheme and the NIHR/Imperial Clinical Research Facility. CI is funded by an NIHR Senior Clinical and Practitioner Research Award.

The Division of Digestive Diseases at Imperial College London receives financial support from the National Institute of Health Research (NIHR) Imperial Biomedical Research Centre (BRC) based at Imperial College London and Imperial College Healthcare NHS Trust.

MW is supported by an Australian National Health and Medical Research Council Investigator Grant (APP1174120).

## Notes

### Competing Interest Statement

The authors have declared no competing interest.

### Funding Statement

This study did not receive any funding

### Author Declarations

The study used (or will use) ONLY openly available human data that were originally located at UK Biobank

